# Third dose of COVID-19 vaccine, CVnCoV, increased neutralizing activity against SARS-CoV-2 wild-type and Delta variant

**DOI:** 10.1101/2022.02.22.22271051

**Authors:** Olaf-Oliver Wolz, Sarah-Katharina Kays, Helga Junker, Sven D. Koch, Philipp Mann, Gianluca Quintini, Philipp von Eisenhart-Rothe, Lidia Oostvogels

**Author notes:** Corresponding author: Dr. Olaf-Oliver Wolz, CureVac AG, Friedrich-Miescher Strasse 15, 72076 Tübingen, Germany.

## Abstract

A third dose of CVnCoV, a former candidate mRNA vaccine against SARS-CoV-2, was previously shown to boost neutralizing antibody responses against SARS-CoV-2 wild-type in adults aged 18-60 and >60 years in a phase 2a clinical study. Here we report neutralizing antibody responses to wild-type and a variant of concern, Delta, after a third dose on day (D) 57 and D180. Neutralization activity was assessed using a microneutralization assay. Comparable levels of neutralizing antibodies against wild-type and Delta were induced. These were higher than those observed after the first two doses, irrespective of age or pre-SARS-CoV-2-exposure status, indicating that the first two doses induced immune memory. Four weeks after the third dose on D180, neutralizing titers for wild-type and Delta were two-fold higher in younger participants than in older participants; seroconversion rates were 100% for wild-type and Delta in the younger group and for Delta in the older group. A third CVnCoV dose induced similar levels of neutralizing responses against wild-type virus and the Delta variant in both naïve and pre-exposed participants, aligning with current knowledge from licensed COVID-19 vaccines that a third dose is beneficial against SARS-CoV-2 variants.

## 1. Introduction

The severe acute respiratory syndrome coronavirus 2 (SARS-CoV-2) that causes coronavirus disease 2019 (COVID-19) is responsible for the current pandemic declared by World Health Organization (WHO) on 11 March 2020 [1]. As of 1 February 2022, there have been nearly 377 million confirmed cases of COVID-19, including more than 5.7 million deaths, reported to WHO and nearly 9.9 billion vaccine doses have been administered worldwide [2]. SARS-CoV-2, an RNA virus, has genetically evolved over time resulting in the emergence of variants from different geographic regions [1]. Some variants with mutations that modify the SARS-CoV-2 spike protein, which is the antigen in authorized COVID-19 vaccines to date, are more resistant to the host neutralization response [3, 4]. The lineage includes three main subtypes (B1.617.1, B.1.617.2 and B.1.617.3), which contain diverse mutations in the receptor-binding domain of the SARS-CoV-2 spike protein. These mutations may increase the immune evasion potential of these variants, although results from some studies suggest cross-neutralization can occur [5-7].

In May 2021 WHO classified the Delta mutant B.1.617.2 as a variant of concern, and in early January 2022 [8], Delta was the most prevalent variant globally [5]. Delta infections can cause typical COVID-19 symptoms but with more severe disease than that caused by wild-type SARS-CoV-2, which is mostly observed in unvaccinated individuals, although some fully vaccinated individuals have also been infected [7].

Recently we reported the results from three clinical trials assessing the safety, efficacy and immunogenicity of a first-generation COVID-19 vaccine candidate: CVnCoV [9-11]. CVnCoV was developed as a sequence-optimized unmodified mRNA vaccine using the proprietary RNActive^®^ technology platform. The mRNA, which encodes a stabilized form of spike-protein from the wild-type strain, is encapsulated in lipid nanoparticles. Clinical development of the first-generation CVnCoV vaccine has been stopped due to the concomitant development of improved second-generation vaccine candidates [12].

Here we report the neutralizing antibody response against the SARS-CoV-2 wild-type and Delta sub-type B.1.617.2 variant after the administration of a third (e.g. booster) dose of CVnCoV vaccine in a subset of the phase 2a trial participants [11].

## 2. Materials and Methods

### 2.1. Study Design and Participants

The methods for this randomized phase 2a clinical trial assessing the safety and immunogenicity of the first generation CVnCoV vaccine have been described previously [11]. In this trial, open-label subsets of participants who had received 12 µg of CVnCoV on Day (D) 1 and D29, received a third dose (12 µg) on D57 (n=30, aged >60 years old) or on D180 (n=30, aged 18-60 years and n=15, aged >60 years).

### 2.2. Assessment of neutralization immune response

Sera were collected before administration of the third dose on D57 and D180, and four weeks later, on D85, and D208, respectively. A microneutralization assay was performed in the laboratories of VisMederi Srl. (Italy) to determine 50% neutralization titers for wild-type and the Delta variant [9]. The wild-type virus was obtained from the European Virus Archive Consortium (Human 2019-nCoV strain 2019-nCoV/Italy-INMI1, clade V; wild type Wuhan strain-like) and the Delta variant (B.1.617.2) was obtained from Institute Pasteur by VisMederi Srl.

Geometric mean titers (GMT) of neutralizing antibodies and their 95% confidence intervals (CI) were determined for the wild-type and Delta SARS-CoV-2 variant and summarized for each time point, according to age group and N-antigen serostatus (see below). Group seroconversion rates for neutralizing antibodies, defined as at least a four-fold increase in titers over baseline and geometric mean-fold rise (GMFR) in titers were calculated from baseline four weeks after the third dose (D85 for the D57 dose and D208 for the D180 dose).

### 2.3. Prior exposure to SARS-CoV-2 virus

To determine if participants were SARS-CoV-2 naïve or pre-exposed, sera were also tested at each time point for antibodies against the SARS-CoV-2 N protein using an ELISA (EI 2606-9601-2 G, EUROIMMUN Medizinische Labordiagnostika AG, Lübeck, Germany), since the vaccine does not contain the N protein.

## 3. Results

### 3.1. Response to third dose in SARS-CoV-2 naïve participants

In the SARS-CoV-2 naïve participants the neutralizing antibody GMTs against wild-type virus and the Delta variant increased on D85 and D208 from the pre-dose levels on D43, following a third dose on D57 and D180, respectively (Figure 1, Table 1). The seroconversion rates and GMFRs were higher after the third doses than after the first two doses (Table 1).

**Table 1.** Geometric mean titers, seroconversion rates and geometric mean fold rises of Delta and wildtype neutralizing antibodies 28 days after two (D43) and three (D85 and D208) CVnCoV doses in SARS-CoV-2 naïve participants

|  | D57 dose >60 years |  | D180 dose >60 years |  | D180 dose 18-60 years |  |
| --- | --- | --- | --- | --- | --- | --- |
|  | Wild-type | Delta | Wild-type | Delta | Wild-type | Delta |
| GMT (95% CI) |  |  |  |  |  |  |
| D43 | 29.5 (16.3-53.5) | 11.8 (6.3-22.1) | 23.8 (7.3-77.5) | 12.3 (3.6-42.6) | 55.6 (26.8-115.2) | 25.9 (12.8-52.5) |
| D85 | 74.6 (46.9-118.9) | 62.8 (36.8-107.2) | - | - | - | - |
| D208 | - | - | 54.6 (23.3-128.4) | 93.5 (52.4-167.0) | 141.7 (101.9-197.1) | 230.2 (148.8-356.1) |
| Seroconversion<sup>a</sup>, n/N (%) |  |  |  |  |  |  |
| D43 | 16/25 (64%) | 5/25 (20%) | 5/10 (50%) | 2/10 (20%) | 15/20 (75%) | 10/20 (50%) |
| D85 | 23/25 (92%) | 20/25 (80%) | - | - | - | - |
| D208 | - | - | 9/10 (90%) | 10/10 (100%) | 20/20 (100%) | 20/20 (100%) |
| GMFR<sup>b</sup> |  |  |  |  |  |  |
| D43 | 5.2 | 2.1 | 4.3 | 2.3 | 9.8 | 3.8 |
| D85 | 13.2 | 11.4 | - | - | - | - |
| D208 | - | - | 9.8 | 17.4 | 25.1 | 33.7 |

<sup>a</sup>Defined as at least four-fold increase in titer over baseline; <sup>b</sup>Geometric mean fold rises of titers over baseline values CI=confidence interval; D=Day; GMFR=geometric mean fold rise; GMT=geometric mean titers; n/N=number of participants of the total number of participants in the subset.

**Figure 1.**
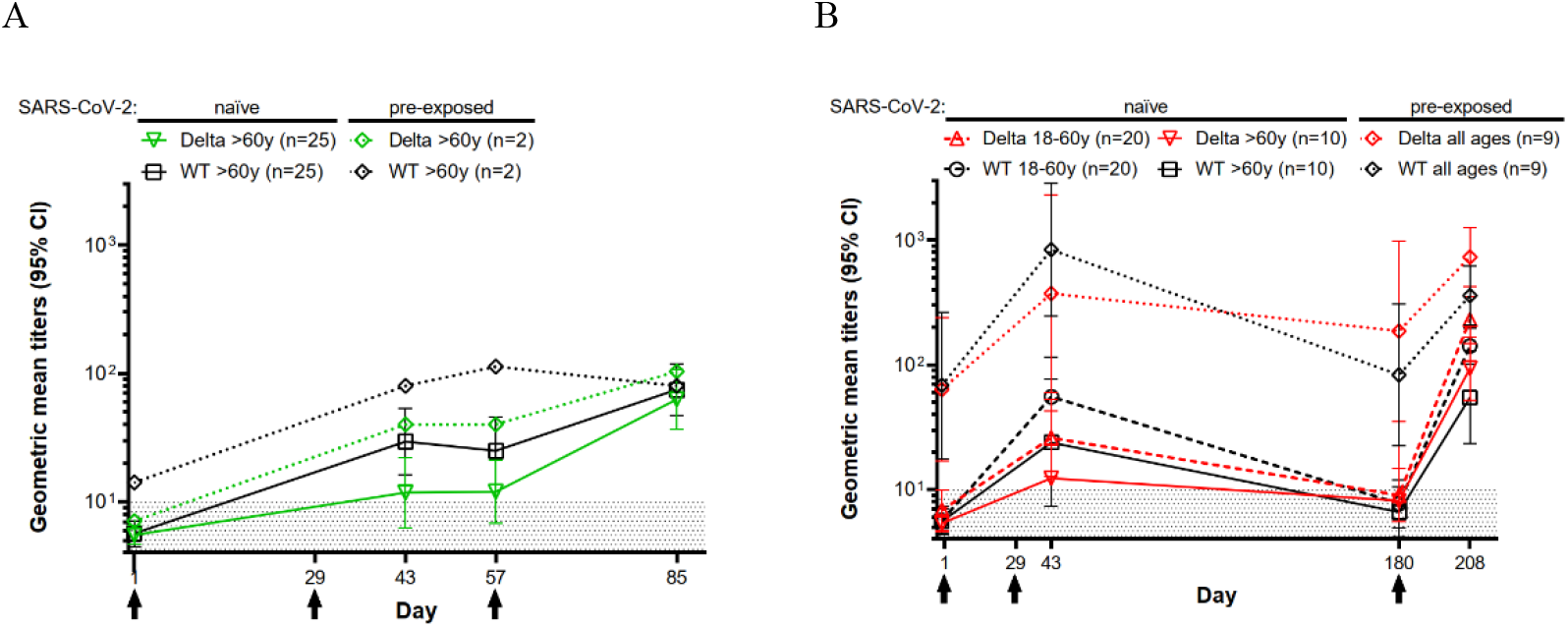
Neutralizing antibody response against wild-type SARS-CoV-2 virus and Delta variant after CVnCoV vaccination. Symbols show geometric mean titers (GMTs) of neutralizing antibodies with 95% confidence intervals (CI) to wild-type (WT, black lines) and Delta (green lines in A and red lines in B) after two and three CVnCoV doses. (A) Participants aged >60 years with a third dose on Day (D) 57 and (B) participants aged 18-60 years and >60 years with a third dose on D180. Solid and dashed lines indicate SARS-CoV-2 naïve participants (seronegative for N protein throughout the trial period) and dotted lines indicate SARS-CoV-2 pre-exposure (seropositive for N protein at baseline). The black arrows indicate the days of vaccinations (D1, D29 (A, B) and D57 (A) or D180 (B)). n = number of participants with available data per group for all time points. Values above the black-dotted region (≥10) were considered positive.

In participants aged >60 years GMTs against both wild-type and Delta on D208 after the third dose on D180 were higher than those on D85 after the third dose on D57 although the 95% CIs overlapped (Figure 1, Table 1). Seroconversion rates were higher after the third dose on D180 than after the third dose on D57 which was also reflected in the higher GMFRs from baseline titers suggesting a better response to the later dose (Table 1).

The GMTs and GMFRs were higher in participants aged 18-60 years than in those >60 years of age after the third dose on D180, although the 95% CIs were overlapping. The seroconversion rates for wild-type were 100% for all participants, irrespective of age, and 100% and 90% for Delta in participants aged 18-60 years and >60 years, respectively (Table 1).

### 3.2. Response to third dose in participants pre-exposed to SARS-CoV-2

Prior to the third dose on D180, the GMTs against wild-type and Delta were still detectable in the nine pre-exposed participants, but not in the naïve participants. On D208 the point estimates for GMTs against wild-type and Delta were higher in the pre-exposed participants than those in naïve participants (Figure 1B). The data for the two pre-exposed participants who received a third dose on D57 have been included for completeness and show a tendency to higher GMTs compared with the naïve participants at the time points shown (Figure 1A).

## 4. Discussion

We recently reported that three doses of CVnCoV (12 µg) had an acceptable safety profile in both young and older adults and induced an increase in IgG and neutralizing antibody titers against the SARS-CoV-2 wild-type [11]. Here we compared the neutralizing antibody responses to the wild-type and Delta variant induced after a third dose of CVnCoV.

Four weeks after the third dose administered on D57 or D180, neutralizing antibody GMTs increased against both the wild-type and Delta variant in SARS-CoV-2 naïve participants above the levels observed on D43 after the first two doses. This demonstrates that the first two doses of CVnCoV had induced immune memory. The neutralizing antibody GMTs against Delta were lower than those against wild-type on D43 after the two doses, but reached similar levels, or higher as those for the wild-type after the third dose, demonstrating a robust immune response against Delta variant was induced. After the D57 dose, neutralizing immune responses against variants of concern, other than Delta, were also increased above those observed after the two doses (data not shown). These findings are consistent with other studies showing that homologous and heterologous mRNA booster vaccines increased immune responses against SARS-CoV-2 variants [13].

In participants aged >60 years, the D180 dose induced higher GMTs (although with overlapping 95% CIs) and seroconversion rates for Delta than the D57 dose. The results suggest that a third dose administered at a later time-point is potentially more immunogenic than the earlier third dose, at least in individuals aged >60 years.

We compared the immune responses to CVnCoV by age because older individuals are at a higher risk of serious SARS-CoV-2 disease. In younger participants GMTs for wild-type and Delta were about two-fold higher than those in adults aged >60 years after two doses of CVnCoV and about 2.5-fold higher after the D180 dose. However, the GMTs were higher after three doses than after two in both younger and older individuals suggesting all participants benefitted from the third dose on D180. Immune responses are known to decline with age due to immunosenescence and this could explain the age-related observations in our study which have also been shown for other COVID-19 vaccines [4, 14].

We previously reported that two doses of CVnCoV induced seroconversion in more than 60% of those who were immunologically naïve for SARS-CoV-2 although GMTs waned to baseline levels on D180 in our phase 1 and 2a studies [7,11]. Here we observed that SARS-CoV-2 pre-exposed individuals still had measurable neutralizing antibodies against both wild-type and Delta up to six months after the first two doses. This indicates that two doses of CVnCoV induced a more potent and longer-lasting immune response against the wild-type and Delta variant in pre-exposed individuals compared with SARS-CoV-2 naïve individuals.

In conclusion, a third CVnCoV dose induced strong neutralizing antibody responses against SARS-CoV-2 wild-type virus in adults aged 18-60 and >60 years demonstrating that the first two doses had induced immune memory. The third dose induced similar levels of neutralizing response against wild-type virus and the Delta variant in both naïve and pre-exposed participants. This is in alignment with the current knowledge from licensed COVID-19 vaccines that a third dose is beneficial against SARS-CoV-2 variants [15-17].

Although the development of CVnCoV has stopped, these cross-neutralizing immune responses against SARS-CoV-2 variants are promising for the next generation SARS-CoV-2 vaccines which are based on the same mRNA platform and are being optimized for variants [18].

## Data Availability

All data produced in the present study are available upon reasonable request to the authors

## Author Contributions

Conceptualization: OOW, SKK, PM, GQ, and LO; Methodology: OOW, SKK, PM, GQ, and LO; Validation: OOW and PvER; Formal Analysis: OOW and SKK; Investigation: OOW, SKK, and HJ; Resources: SDK and GQ; Writing – Original Draft Preparation: OOW, SKK, HJ, SDK, PM, and GQ; Writing – Review & Editing: OOW, SKK, HJ, SDK, PM, GQ, PvER, and LO; Visualization: OOW and SKK; Supervision: HJ, SDK, PM, GQ and LO; Project Administration: HJ, PM and LO.

All authors have read and agreed to the published version of the manuscript.

## Institutional Review Board Statement

The study was conducted in accordance with the Declaration of Helsinki, and the research protocol was approved by the Institutional Review Board of Peru and Panama. It is registered at ClinicalTrials.gov (NCT04515147).

## Informed Consent Statement

Informed consent was obtained from all subjects involved in the study.

## Data Availability Statement

The data presented in this study are available on request from the corresponding author. The data are not publicly available due to [insert reason here].

## Acknowledgments

The authors are grateful to Giulia Lapini, Alessandro Torelli, Emanuele Montomoli and laboratory team from VisMederi Srl for their continued support and commitment in performing the neutralizing antibody assays. We thank Dominik Vahrenhorst, CureVac Bioinformatics, for analytical support, Giulia Povellato and Robert Tensen, Oliver Schönborn-Kellenberger for helpful comments on manuscript drafts, and Merel Hazewindus and Margaret Haugh for medical writing and editorial services.

## References

1. WHO. Available online: https://www.who.int/en/activities/tracking-SARS-CoV-2-variants/ (accessed on 2 February 2022).

2. WHO. Available online: https://covid19.who.int/ (accessed on 2 February 2022).

3. Wibmer CK, Ayres F, Hermanus T, Madzivhandila M, Kgagudi P, Oosthuysen B, Lambson BE, de Oliveira T, Vermeulen M, van der Berg K, Rossouw T, Boswell M, Ueckermann V, Meiring S, von Gottberg A, Cohen C, Morris L, Bhiman JN, Moore PL. SARS-CoV-2 501Y.V2 escapes neutralization by South African COVID-19 donor plasma. Nat Med. 2021, 27, 622–625; DOI: 10.1038/s41591-021-01285-x.

4. Wang J, Tong Y, Li D, Li J, Li Y. The impact of age difference on the efficacy and safety of COVID-19 vaccines: a systematic review and meta-analysis. Front Immunol. 2021, 6, 12, 758294; DOI: 10.3389/fimmu.2021.758294.

5. Planas D, Veyer D, Baidaliuk A, Staropoli I, Guivel-Benhassine F, Rajah MM, Planchais C, Porrot F, Robillard N, Puech J, Prot M, Gallais F, Gantner P, Velay A, Le Guen J, Kassis-Chikhani N, Edriss D, Belec L, Seve A, Courtellemont L, Péré H, Hocqueloux L, Fafi-Kremer S, Prazuck T, Mouquet H, Bruel T, Simon-Lorière E, Rey FA, Schwartz O. Reduced sensitivity of SARS-CoV-2 variant Delta to antibody neutralization. Nature. 2021, 596, 276–280; DOI: 10.1038/s41586-021-03777-9.

6. Mileto D, Fenizia C, Cutrera M, Gagliardi G, Gigantiello A, De Silvestri A, Rizzo A, Mancon A, Bianchi M, De Poli F, Cuomo M, Burgo I, Longo M, Rimoldi SG, Pagani C, Grosso S, Micheli V, Rizzardini G, Grande R, Biasin M, Gismondo MR, Lombardi A. SARS-CoV-2 mRNA vaccine BNT162b2 triggers a consistent cross-variant humoral and cellular response. Emerg Microbes Infect. 2021, 10, 2235–2243; DOI: 10.1080/22221751.2021.2004866.

7. Bian L, Gao Q, Gao F, Wang Q, He Q, Wu X, Mao Q, Xu M, Liang Z. Impact of the Delta variant on vaccine efficacy and response strategies. Expert Rev Vaccines. 2021, 20, 1201–1209; DOI: 10.1080/22221751.2021.2004866.

8. WHO Available online: https://www.cdc.gov/coronavirus/2019-ncov/variants/delta-variant.html (accessed on 2 February 2022)

9. Kremsner PG, Mann P, Kroidl A, Leroux-Roels I, Schindler C, Gabor JJ, Schunk M, Leroux-Roels G, Bosch JJ, Fendel R, Kreidenweiss A, Velavan TP, Fotin-Mleczek M, Mueller SO, Quintini G, Schönborn-Kellenberger O, Vahrenhorst D, Verstraeten T, Alves de Mesquita M, Walz L, Wolz OO, Oostvogels L; CV-NCOV-001 Study Group. Safety and immunogenicity of an mRNA-lipid nanoparticle vaccine candidate against SARS-CoV-2: A phase 1 randomized clinical trial. Wien Klin Wochenschr. 2021, 133, 931–941; DOI: 10.1080/22221751.2021.2004866.

10. Kremsner PG, Ahuad Guerrero RA, Arana-Arri E, Aroca Martinez GJ, Bonten M, Chandler R, Corral G, De Block EJL, Ecker L, Gabor JJ, Garcia Lopez CA, Gonzales L, Granados González MA, Gorini N, Grobusch MP, Hrabar AD, Junker H, Kimura A, Lanata CF, Lehmann C, Leroux-Roels I, Mann P, Martinez-Reséndez MF, Ochoa TJ, Poy CA, Reyes Fentanes MJ, Rivera Mejia LM, Ruiz Herrera VV, Sáez-Llorens X, Schönborn-Kellenberger O, Schunk M, Sierra Garcia A, Vergara I, Verstraeten T, Vico M, Oostvogels L; HERALD Study Group. Efficacy and safety of the CVnCoV SARS-CoV-2 mRNA vaccine candidate in ten countries in Europe and Latin America (HERALD): a randomised, observer-blinded, placebo-controlled, phase 2b/3 trial. Lancet Infect Dis. 2021, 23, S1473-3099(21)00677-0; DOI: 10.1080/22221751.2021.2004866.

11. Sáez-Llorens X, Lanata C, Aranguren E, Celis CR, Cornejo R, DeAntonio R, Ecker L, Garrido D, Gil AI, Gonzales M, Hess-Holtz M, Leroux-Roels G, Junker H, Kays SK, Koch SD, Lazzaro S, Mann P, Quintini G, Srivastava B, Vahrenhorst D, Von Eisenhart-Rothe P, Wolz OO, Oostvogels L. Safety and immunogenicity of mRNA-LNP COVID-19 vaccine CVnCoV in Latin American adults; a phase 2 randomized study. Vaccine X. submitted.

12. CureVac AG. Available online: https://www.curevac.com/en/2021/10/12/curevac-to-shift-focus-of-covid-19-vaccine-development-to-second-generation-mrna-technology/ (Accessed on 2 February 2022).

13. Munro APS, Janani L, Cornelius V, Aley PK, Babbage G, Baxter D, Bula M, Cathie K, Chatterjee K, Dodd K, Enever Y, Gokani K, Goodman AL, Green CA, Harndahl L, Haughney J, Hicks A, van der Klaauw AA, Kwok J, Lambe T, Libri V, Llewelyn MJ, McGregor AC, Minassian AM, Moore P, Mughal M, Mujadidi YF, Murira J, Osanlou O, Osanlou R, Owens DR, Pacurar M, Palfreeman A, Pan D, Rampling T, Regan K, Saich S, Salkeld J, Saralaya D, Sharma S, Sheridan R, Sturdy A, Thomson EC, Todd S, Twelves C, Read RC, Charlton S, Hallis B, Ramsay M, Andrews N, Nguyen-Van-Tam JS, Snape MD, Liu X, Faust SN; COV-BOOST study group. Safety and immunogenicity of seven COVID-19 vaccines as a third dose (booster) following two doses of ChAdOx1 nCov-19 or BNT162b2 in the UK (COV-BOOST): a blinded, multicentre, randomised, controlled, phase 2 trial. Lancet. 2021, 398, 2258–2276; DOI: 10.1016/S0140-6736(21)02717-3.

14. Müller L, Andrée M, Moskorz W, Drexler I, Walotka L, Grothmann R, Ptok J, Hillebrandt J, Ritchie A, Rabl D, Ostermann PN, Robitzsch R, Hauka S, Walker A, Menne C, Grutza R, Timm J, Adams O, Schaal H. Age-dependent immune response to the Biontech/Pfizer BNT162b2 coronavirus disease 2019 vaccination. Clin Infect Dis. 2021, 6; 73, 2065–2072; DOI: 10.1080/22221751.2021.2004866.

15. Choi A, Koch M, Wu K, Chu L, Ma L, Hill A, Nunna N, Huang W, Oestreicher J, Colpitts T, Bennett H, Legault H, Paila Y, Nestorova B, Ding B, Montefiori D, Pajon R, Miller JM, Leav B, Carfi A, McPhee R, Edwards DK. Safety and immunogenicity of SARS-CoV-2 variant mRNA vaccine boosters in healthy adults: an interim analysis. Nat Med. 2021, 27, 2025–2031; DOI: 10.1038/s41591-021-01527-y.

16. Falsey AR, Frenck RW Jr, Walsh EE, Kitchin N, Absalon J, Gurtman A, Lockhart S, Bailey R, Swanson KA, Xu X, Koury K, Kalina W, Cooper D, Zou J, Xie X, Xia H, Türeci Ö, Lagkadinou E, Tompkins KR, Shi PY, Jansen KU, Şahin U, Dormitzer PR, Gruber WC. SARS-CoV-2 neutralization with BNT162b2 vaccine dose 3. N Engl J Med. 2021, 385, 1627–1629; DOI: 10.1056/NEJMc2113468.

17. Cromer D, Steain M, Reynaldi A, Schlub TE, Wheatley AK, Juno JA, Kent SJ, Triccas JA, Khoury DS, Davenport MP. Neutralising antibody titres as predictors of protection against SARS-CoV-2 variants and the impact of boosting: a meta-analysis. Lancet Microbe. 2022, 3, e52–e61; DOI: 10.1016/S2666-5247(21)00267-6.

18. Gebre MS, Rauch S, Roth N, Yu J, Chandrashekar A, Mercado NB, He X, Liu J, McMahan K, Martinot A, Martinez DR, Giffin V, Hope D, Patel S, Sellers D, Sanborn O, Barrett J, Liu X, Cole AC, Pessaint L, Valentin D, Flinchbaugh Z, Yalley-Ogunro J, Muench J, Brown R, Cook A, Teow E, Andersen H, Lewis MG, Boon ACM, Baric RS, Mueller SO, Petsch B, Barouch DH. Optimization of non-coding regions for a non-modified mRNA COVID-19 vaccine. Nature. 2022, 601, 410–414; DOI: 10.1038/s41586-021-04231-6.

